# Prospective SARS-CoV-2 cohort study among general practitioners during the second COVID-19 wave in Flanders, Belgium

**DOI:** 10.1101/2021.03.26.21254327

**Authors:** Joachim Mariën, Ann Ceulemans, Diana Bakokimi, Christine Lammens, Margareta Ieven, Stefan Heytens, An De Sutter, Jan Y Verbakel, Ann Van den Bruel, Herman Goossens, Pierre Van Damme, Kevin K. Ariën, Samuel Coenen

**Author notes:** Corresponding author: Samuel Coenen, Campus Drie Eiken, Gouverneur Kinsbergencentrum, Doornstraat 331, 2610 Antwerp (wilrijk), Belgium.

## Abstract

Primary health care providers (PHCPs), especially general practitioners (GPs) are essential to organise health care efficiently. During the COVID-19 pandemic, they also keep the pressure off hospitals. PHCPs are assumed to be at high risk of a COVID-19 infection, as they are exposed to a large portion of the population (usually with less personal protective equipment than other frontline health care workers(HCWs)). Nevertheless, previous seroprevalence studies focussed on the general population or HCWs in hospital settings, rather than PHCPs. The aim of this study was to determine the seroprevalence of PHCPs after the first and during the second SARS-CoV-2 wave in Flanders (Belgium) and compare it to the seroprevalence in the general population (blood donors). A prospective cohort of PHCPs, mainly GPs (*n*=698) was screened for IgG antibodies against SARS-CoV-2 at five different time-points (June-December 2020). The dried blood spots they produced were analysed using a Luminex multiplex immunoassay. The seroprevalence of PHCPs remained stable between June and September 2020 (4.6-5.0%), but increased significantly from October to December (8.1-13.4%) 2020. The seroprevalence of PHCPs was not significantly higher than the seroprevalence of the blood donors at the end of December 2020. In conclusion, the sharp increase in seroprevalence during the second COVID-19 wave in Flanders shows that PHCPs were more at risk during the second wave compared to the first one. However, the increase was in line with the general population suggesting that PHCPs mainly got infected in their private settings.

## Introduction

In 2020, Coronavirus disease 19 (COVID-19) affected many European countries in a first wave that occurred in spring and declined after far-reaching lockdowns, and a second wave that emerged in autumn (1). Belgium has been hit particularly hard by this second pandemic wave and even had the highest per capita case numbers in Europe during its epidemic peak (2). While these waves are typically monitored in terms of PCR-confirmed cases, serosurveillance data represent the accumulative number of infections and suggests many more undiagnosed cases.

High seroprevalence rates are typically observed among primary health care providers (PHCPs), as they manage the vast majority of symptomatic COVID-19 patients (3). Among the PHCPs, general practitioners (GPs) are essential to organise health care efficiently given their role as gatekeepers to the next levels of care by keeping pressure off hospitals and triaging patients (4). If primary care cannot be delivered safely, the COVID-19 epidemic disrupts general public health by failing to deliver non-COVID-19 related healthcare, exemplified by the collapse of the health system in northern Italy during the first wave where 38% of the physicians who died from COVID-19 were actually GPs (5). Furthermore, asymptomatically infected GPs might play a crucial role in the transmission of COVID-19 to their patients as a vector or as a source of transmission themselves (6). Although GPs are typically not involved in aerosol generating procedures (e.g. in comparison to pulmonologists), they do bear the brunt of an epidemic from frequent contact with a very large population in less secure circumstances. Therefore, preserving the capacity of GPs and their co-workers throughout the COVID-19 epidemic is essential. In Belgium, this is particularly the case as the GP workforce consists of older adults (mean age = 55) and is therefore at higher risk for COVID-19 morbidity and mortality (7). However, while many serological surveys have been performed on health care workers (HCWs) in hospital settings, surprisingly little surveys followed PHCPs over time (3). Here, we evaluated the SARS-CoV-2 seroprevalence rate in a prospective cohort of PHCPs after the first and during the second wave in Flanders (the largest of the three regions in Belgium).

## Method

### Study design and setting

In May 2020, we recruited PHCPs that are currently working in a general practice and physically manage patients/clients in Flanders. Participants that provided written informed consent were asked to self-sample capillary blood by fingerpicking and to complete a baseline questionnaire through an online secured application (basic socio-demographics, health status, the availability and implementation of preventive measures) at five different time-points (June-September-October-November and December 2020). The blood samples were stored on Whatman903 protein saver card and returned via regular mail to the University of Antwerp, where they were preserved in the dark at 4°C.

### Laboratory

All dried blood spots (DBS) were analysed for the presence of anti-SARS-CoV-2 IgG antibodies at the Institute of Tropical Medical in Antwerp using an in-house Luminex multiplex immunoassay (MIA), which is described in detail in Mariën et al 2020 (8). In brief, we punched 2 discs (4mm) from each DBS in an Eppendorf tube and added dilution buffer (hypertonic PBS buffer) to a final concentration of 1/600. The dilutions were mixed with 1.25×10^6^ paramagnetic MAGPLEX COOH-microspheres from Luminex Corporation (Texas, USA) that were coupled with recombinant receptor binding domain, whole spike protein and nucleocapsid protein (BIOCONNECT, The Netherlands). After incubation of beads and diluted sera, biotin-labelled anti-human IgG (1:125) and streptavidin-R-phycoerythrin (1:1000) conjugate were added. The beads were read using a Luminex® Bio-Plex 100/200 analyser and expressed as signal-to-noise ratios.

### Data analysis

Samples were considered to be positive if the fluorescent signal was more than 2 times the standard deviation + mean of negative controls (*n*=96) for all antigens, which corresponds to a specificity of 99% (8). Samples that surpassed the cut-off values for only two antigens (and if the signal of the third antigen was higher than the mean of negative control) were considered to be uncertain. Empty serological time points (when a participant did not return the DBS at a particular sampling time-point) were filled based on previous and later serological results if appropriate. We also considered all seroreverted samples to be positive at later time points, as it is known that most people develop a long-lasting antibody immunity (at least 6 months after infection). Only positive samples were considered for final seroprevalence estimates. To put our data in perspective (Fig 1), we also included seroprevalence estimates from blood donors (representing the general population) that were obtained from surveillance studies at the University of Antwerp (June and September; 9), and Sciensano and Rode Kruis Vlaanderen (November and December; 10). These samples were screened on SARS-Cov-2 IgG antibodies using the EuroImmun (June and September) or Wantai (November-December) SARS-CoV-2 antibody ELISAs.

**Fig 1:**
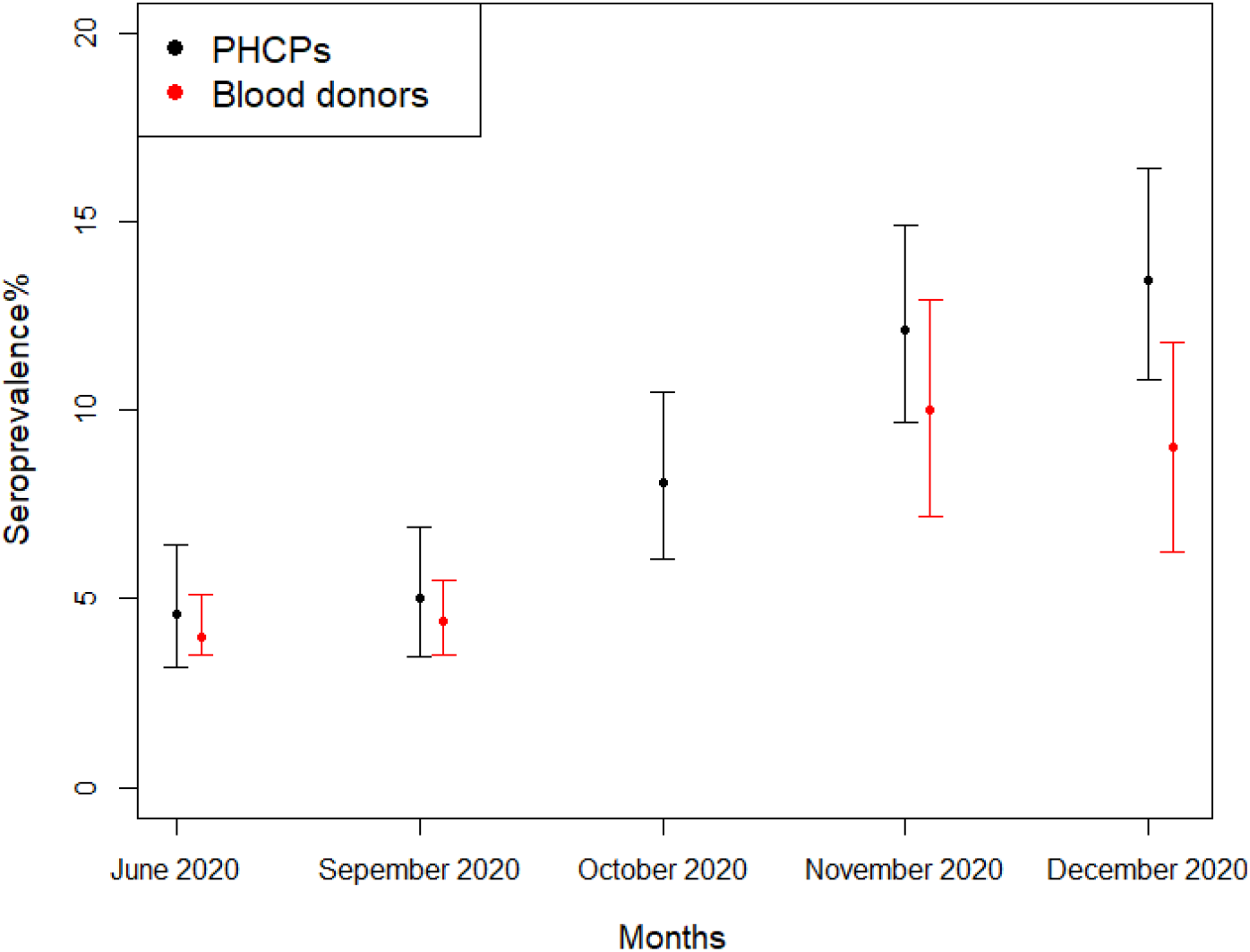
SARS-CoV-2 seroprevalence in a cohort of primary health care providers (PHCPs) after the first and during the second COVID-19 wave in Flanders in 2020 (black) at the end of each month. Blood donor data were obtained from serosurveillance studies at the University of Antwerp (June and September; 9), and Sciensano and Rode Kruis Vlaanderen (November and December; 10).

## Results

From all the PHCPs that responded to our call to participate in the study, 698 provided informed consent and returned at least one DBS to the University of Antwerp. The willingness to participate remained high throughout the study, as the majority of participant (*n*=604) returned a DBS at more than three time points. The cohort of participants represents a convenience sample of PHCPs, mainly GPs (n=641), women (n=520) and a mean age of 41.1 years (SD=11.6) (Table 1). The seroprevalence of PHCPs after the first COVID-19 wave was 4.6% (95%CI 3.2-6.4%) in June, which is in line with the seroprevalence of blood donors (4.9%; 95%CI 3.1-5.9%), (9) but lower than the seroprevalence of hospital HCWs (9.4%; 95%CI 6.5-13.4%) (11) at that time in Flanders. The seroprevalence of PHCPs remained stable until September, but increased significantly from October to December to 13.4% (95%CI 10.8-16.4%). The highest increase (Δ4%) was found between October and November, which coincided with the peak in confirmed PCR cases in the community. At the end of the second wave (December 2020), the seroprevalence of PHCPs (13.4%, 95%CI 10.8-16.4%) was not significantly higher than the seroprevalence of blood donors (9.0%, 95% CI 6.2-11.8%, personal communication R. De Pauw Sciensano) (10) (Fig 1).

**Table 1.** Characteristics of primary healthcare providers (PHCPs), including general practitioners (GPs) and other PHCPs who participated in at least one testing time point.

|  | All PHCPs<br>n=698 |  | GPs<br>n=641 |  | Other PHCPs<br>n=57 |  |
| --- | --- | --- | --- | --- | --- | --- |
| Age, mean (SD) | 41 (12) |  | 41 (12) |  | 40 (11) |  |
| Gender<sup>(1)</sup>, n (%) |  |  |  |  |  |  |
| - Male | 173 | (24.8) | 166 | (25.9) | 7 | (12.3) |
| - Female | 520 | (74.5) | 470 | (73.3) | 50 | (87.7) |
| - Not reported | 5 | (0.7) | 5 | (0.8) |  |  |
| Practice size, n (%)<sup>(1)</sup> |  |  |  |  |  |  |
| - Solo | 84 | (19.7) | 84 | (19.9) | 3 | (7.3) |
| - Duo | 83 | (19.4) | 83 | (19.6) | 3 | (7.3) |
| - Group (<8 employees) | 139 | (32.6) | 138 | (32.6) | 6 | (14.6) |
| - Big group (>7) | 112 | (26.2) | 111 | (26.2) | 27 | (65.85) |

<sup>(1)</sup> if numbers do not add up to the column total, this is due to missing data; numbers of practices for PHCPs=427, GPs=423 and other PHCPs=41.

## Discussion

This cohort study shows a significant increase of antibodies against SARS-CoV-2 in DBS samples between the end of the first and end of the second wave in Flanders, but did not find a significant difference in seroprevalence between PHCPS and the general population (blood donors). We assume that the small difference in seroprevalence of PHCPs compared to the general population in December (4.4% higher in PHCPS) reflects a true difference, as DBS leads to a decrease in sensitivity compared to serum meaning the actual difference might be higher (12). Indeed, a limitation of our study is that we could not directly compare the seroprevalence of PHCPs to blood donors because samples were different (capillary vs venous blood), stored differently (DBS vs serum tube) and analysed with different techniques (Luminex MIA versus ELISA). We chose DBS sampling because of its convenience of non-invasive sampling and room-temperature storage. Serological point-of-care tests (POCTs) would have substantially improved the timeliness of the test results and the PHCPs could have immediately checked their results (13). We did not use POCTs in this study because of limited resources and particularly since their specificity was not good enough at the time we conceived this study. Nonetheless, our study suggests that DBS sampling is a useful alternative for SARS-CoV-2 serosurveillance in nonclinical, resource-limited settings. Samples can also be reused (e.g. to check for co-infections), which is impossible with POCTs.

In conclusion, the steep increase in seroprevalence during the second COVID-19 wave indicates that PHCPs were more at risk during the second wave compared to the first wave in Flanders. Although we could not verify if an infection occurred at work or at home, the slightly higher incidence (insignificant) of PHCPs compared to blood donors suggests that only a minority of GPs became infected when managing patients. This suggests that GPs were relatively well protected during the second COVID-19 wave or that close contacts with infected patients were limited.

## Data Availability

The datasets used and/or analysed during the current study are available from the corresponding author on reasonable request.

## Ethics statement

Ethical approval for the study was given by the institutional review board of the Antwerp University Hospital/University of Antwerp (20/24/315). The study was also registered as a clinical trial (ClinicalTrials.gov Identifier: NCT04779424). All participants provided informed consent to participate. We declare that the planning conduct and reporting of the study was in line with the Declaration of Helsinki, as revised in 2013.

## Acknowledgements

We are grateful for the support by Domus Medica and the academic centres of general practice of UAntwerp, UGhent and KULeuven for the recruitment, for the help of all those making the study materials available to the study participants and for the altruistic contribution of the latter. We are also grateful for the support of this study by Sereina Herzog, Elza Duysburgh, Isabelle Desombere, Robby de Pauw and Magali Le Clef.

## Funding

The work was funded by the University of Antwerp (BOF/COVID 42828), the RECOVER project (European Union’s Horizon 2020 research and innovation programme; No 101003589), the Research Foundation Flanders (FWO) (G0G4220N and G054820N), the Health Care Worker seroprevalence study (Sciensano/ITM), NCT04373889 and intramural funds from the Institute of Tropical Medicine Antwerp. Joachim Mariën is currently a research assistant of Research Foundation Flanders (FWO).

## Competing interests

The funder of the study had no role in the study design; collection, management, analysis, or interpretation of the data, preparation, review, or approval of the manuscript; or decision to submit the manuscript for publication.

## Author statement

Conceived the study: SC. Wrote the paper: JM & SC. Performed the lab experiments: JM, AC, DB. Performed the data analyses: JM & SC. Supervised data collection and laboratory work: JM, CL, GL, SH, AdS, JV, AvdB, HG, PcD, SC, KA. All authors read and approved the final manuscript.

## Notes

### Competing Interest Statement

The authors have declared no competing interest.

### Clinical Trial

ClinicalTrials.gov Identifier: NCT04779424

### Clinical Protocols

https://www.clinicaltrials.gov/ct2/show/NCT04779424

### Funding Statement

The work was funded by the University of Antwerp (BOF/COVID 42828), the RECOVER project (European Unions Horizon 2020 research and innovation programme; No 101003589), the Research Foundation Flanders (FWO) (G0G4220N and G054820N), the Health Care Worker seroprevalence study (Sciensano/ITM), NCT04373889 and intramural funds from the Institute of Tropical Medicine Antwerp. Joachim Marien is currently a research assistant of Research Foundation Flanders (FWO).

### Author Declarations

Ethical approval for the study was given by the institutional review board of the Antwerp University Hospital/University of Antwerp (20/24/315). The study was registered as a clinical trial (ClinicalTrials.gov Identifier: NCT04779424) All participants provided informed consent to participate. We declare that the planning conduct and reporting of the study was in line with the Declaration of Helsinki, as revised in 2013.

